# Trends of health and dietary disparities by economic status among elderly individuals in Japan from 2004 to 2014: A repeated cross-sectional survey

**DOI:** 10.1101/2023.04.24.23289068

**Authors:** Daisuke Machida

## Abstract

This study examines the trends in health and dietary disparities by economic status among elderly individuals in Japan from 2004 to 2014 with subjective measures. The study design utilized a repeated cross-sectional approach, using data from the Survey of Attitudes among the Elderly toward Daily Life in 2004 and 2014. Logistic regression analysis was performed with subjective economic status, survey year, and their interactions as independent variables, and self-rated health, dietary satisfaction, and intake of balanced meals as dependent variables. The results revealed that disparities in self-rated health, dietary satisfaction, and intake of a balanced meal were present due to economic status. Furthermore, the disparities in self-rated health, dietary satisfaction, and balanced meal intake by economic status remained unchanged from 2004 to 2014 (p for interaction ≥ 0.05). The findings were consistent in sensitivity analyses conducted on those aged 75 and older, as well as on long-term care insurance recipients.

## 1. Introduction

Economic inequality affects people’s health and dietary behaviors. A meta-analysis of multilevel studies showed that economic inequality increases the relative risk for mortality from prospective cohort studies and the odds ratio for poor self-rated health from cross-sectional studies [1]. Additionally, a review article exploring the causal relationship between income inequality and health using a causal inference framework concluded that a causal relationship exists wherein income inequality affects health [2]. Economic inequality affects health through unhealthy behaviors [3], and unhealthy eating habits is one of them. In fact, good socioeconomic status is associated with high consumption of whole grains, vegetables and fruits, lean meats, and seafood; conversely, poor socioeconomic status is associated with high consumption of refined grains, potatoes, fatty meats, and fried foods [4]. Moreover, poor socioeconomic status is associated with obesity [5]. These mechanisms include the fact that foods that cost little money or time have high energy density and low nutrient density [4,5,6].

Health disparities by economic status are known in Japan. The National Health and Nutrition Survey conducted by Japan’s Ministry of Health, Labour and Welfare investigates disparities in health and diet by income among adults and the elderly every few years in the 2010s [7–11]. Eating habits, physique, and health habits vary by income, and this has remained generally unchanged in the 2010s [7–10,12]. The National Health and Nutrition Survey began capturing data on health disparities by income in the 2010s, and changes from earlier years are unavailable. However, in Japan, trends in health inequalities by socioeconomic status have been captured since 1986 using data from the Comprehensive Survey of Living Conditions [13–17]. One study, which evaluated long-term trends from 1986 to 2013, found that health inequalities by income are cyclical [13]. This makes sense because Japan’s economy fluctuated wildly from the 1990s through the 2010s. Following the collapse of Japan’s bubble economy in the 1990s, the country experienced a prolonged period of economic stagnation. Despite some recovery in 2000, the economy was significantly impacted by the Lehman shock in 2009 and the Great East Japan Earthquake in 2011. This fluctuation in Japan’s economic situation can be clearly seen in the unemployment rate [13]. The impact of each economic change on health and diet is also analyzed and includes dietary changes due to the Lehman shock [18], health outcomes during the employment ice age [19], and changes in health disparities due to the collapse of the bubble economy [14].

Japan has one of the fastest-aging populations in the world [20]. As population aging is expected to continue in developed countries worldwide, conducting research on health promotion targeting Japan’s elderly will provide valuable evidence for future health promotion in these countries. The same can be said about the issue of disparities in health and diet. Several studies have examined health and diet disparities by economic status among Japan’s elderly, confirming the existence of disparities in health and diet by economic status [21–24]. Additionally, trends in health disparities by income among Japan’s elderly have been examined [25]. However, trends in dietary disparities by economic status among the elderly have not been explored.

In this study, I describe changes in health dietary disparities owing to economic conditions from 2004 to 2014 for Japan’s elderly population. I hypothesized that changes in the situations of health disparities before and after this period will occur as this period was interspersed with periods of significant economic volatility, including the Lehman Shock and Great East Japan Earthquake.

## 2. Materials and Methods

### 2.1. Study Design and Data

This study used a repeated cross-sectional survey design. Data were derived from the Survey of Attitudes among the Elderly toward Daily Life 2004 and 2014 (Director General for Policies on Cohesive Society, Cabinet Office, The Government of Japan) and obtained from the Social Science Japan Data Archive, Center for Social Research and Data Archive [26–28]. The survey was conducted to determine the actual conditions and attitudes of the elderly regarding their overall daily life. This includes information on their daily living conditions, life satisfaction, food, clothing, and housing, and satisfaction with household chores, going out, daily enjoyment, and information about daily life. The survey targeted persons aged 60 and over throughout Japan and has been conducted four times so far in 1998, 2004, 2009, and 2014. Data from the 2004 and 2014 surveys are available by applying to the Social Science Japan Data Archive, Center for Social Research and Data Archive, and were used in a secondary analysis. The survey was conducted from November to December and was conducted through interview and mail in 2004 and 2014, respectively. In 2004, 4000 people were surveyed, with 2862 (71.6%) valid responses. In 2014, 6000 people were surveyed, with 3893 (64.9%) valid responses.

This study was conducted using anonymous information from a previously completed survey and according to the ethical guidelines for life science and medical research involving human subjects in Japan [29].

### 2.2. Variables

#### 2.2.1. Economic status

I used subjective economic status measured by a single question in this study. Participants were asked, “How would you describe your current economic situation?.” The following five options were then given: “I have a comfortable household budget and am not worried at all,” “I do not have a very comfortable budget, but I am not too worried,” “I do not have a comfortable budget, and I am somewhat worried,” “I have a very tight budget, and I am very worried,” and “I don’t know [26,27].” I excluded the last option from the analyses. I divided the options into “no economic insecurity (ref.) (sum of the first two options) “and “have economic insecurity (sum of the latter two options)” and were used in the analyses.

In Japan, income declines as many people leave the workforce at ages 60–65. Although some income can be obtained from public pensions, this is often less than the salary income up to that point. In fact, the average income of elderly households is less than half the average income of nonelderly households [30]. Conversely, the amount of savings is higher among the elderly [31]. Economic comfort in the lives of Japan’s elderly largely depends on their savings funds. Therefore, income alone is insufficient for understanding the elderly’s economic situation in Japan. Subjective economic status has also been shown to correlate with income among Japanese adults, making it an appropriate indicator of comprehensive economic status, including income [32]. For these reasons, this study adopted subjective economic status as an indicator.

#### 2.2.2. Outcomes

I used self-rated health, dietary satisfaction, and balanced meal intake as outcomes.

The participants were asked to rate their self-rated health on a five-point scale: good, somewhat good, fair, somewhat poor, and poor. In this study, the categories “good/fair (ref.) (sum of good, somewhat good, and fair)” and “poor (sum of somewhat poor and poor)” were used for analysis.

The participants were given the question, “Are you satisfied with your overall diet?” a one-choice answer from the following five options: satisfied, somewhat satisfied, somewhat dissatisfied, dissatisfied, and don’t know. Dietary satisfaction is a representative construct of diet-related quality of life [33,34]. In this study, this response was divided into “satisfied (ref.) (sum of satisfied and somewhat satisfied)” and “dissatisfied (sum of somewhat dissatisfied and dissatisfied)” for analysis. Responses of “don’t know” were treated as missing values.

To assess balanced meal intake, participants were asked, “What are your concerns regarding your daily diet?” The response option “lack of dietary balance” was included, and answers to this question were used. Participants who selected this option were considered to have said “no.” Those who did not select this option were considered to have said “yes (ref.).” A well-balanced meal intake is associated with lower mortality rates [35,36] and is therefore an appropriate indicator of a healthy diet.

#### 2.2.3. Other

In addition to the above, was used the survey year (2004(ref.), 2014), gender (men, women), age (60–69, 70–79, ≥80), living arrangements (living together, living alone), and long-term care insurance recipient.

### 2.3. Analysis

I used responses from 6587 individuals (2004y: 2832, 2014y: 3755) with no missing required items for the analyses. For analyses regarding dietary satisfaction, I excluded responses containing an additional 107 missing values from the above in 2014.

First, I described participant characteristics for each survey year and performed chi-square tests. I then described the number and percentage of participants in each year for each economic status in terms of self-rated health, dietary satisfaction, and balanced diet intake. This was not only captured for the all participants but also for those aged 75 and older only for long-term care insurance recipients only. Then, I used logistic regression analysis to determine the main effects of economic status and survey year, and identify temporal changes in outcomes using interaction of economic status and survey year. In doing so, economic status, survey year, and their interaction were used as independent variables in Model 1. In Model 2, I adjusted for gender, age, and living arrangements. In addition to the overall analysis, I conducted sensitivity analyses among those who are aged 75 and older, and those who are long-term care insurance recipients.

All analyses were conducted using IBM SPSS Statistics 28.0, with a 5% significance level (two-tailed test). Python 3.9.7 was used to create the figures.

## 3. Results

### 3.1. Participants’ Characteristics

**Table 1** shows the participants’ characteristics according to survey years. Age (p < 0.001), living arrangements (p < 0.001), economic status (p < 0.001), and dietary satisfaction (p < 0.001) were significantly different by survey years. Additionally, the ratios of people aged 75 or older (p < 0.001) and long-term care insurance recipients (p < 0.001) also differed by survey years.

**Table 1:**
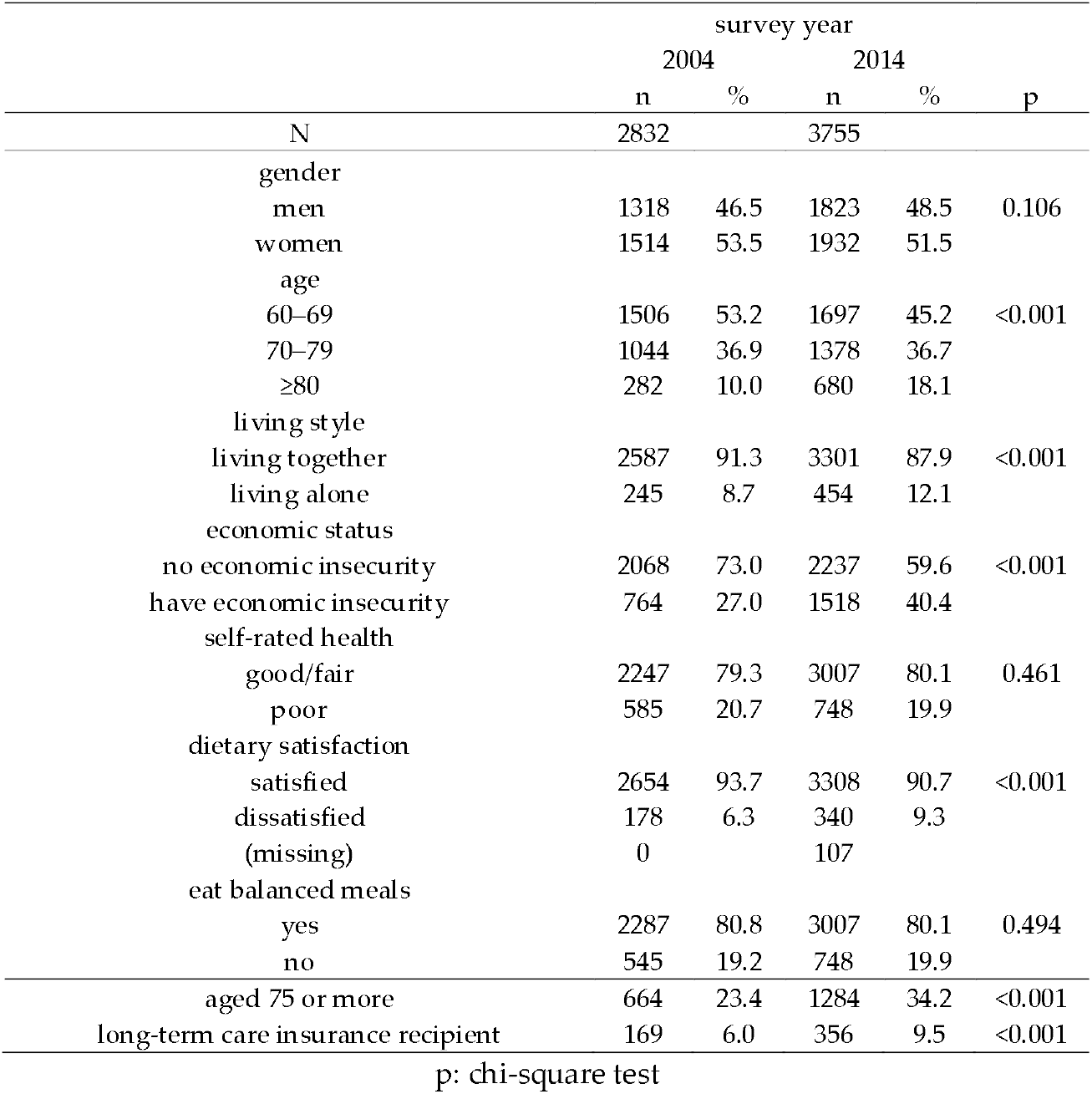
Participants’ characteristics according to survey years

### 3.2. Self-rated health, dietary Satisfaction, and Balanced Meals Intake for Each Year by Economic Status

**Table 2** shows self-rated health, dietary satisfaction, and balanced meal intake for each year by economic status. For economically insecure participants, the percentage of those dissatisfied with the general diet was 13.9% and 16.8% in 2004 and 2014, respectively. In contrast, for the participants who did not have economic insecurity, the percentage of those dissatisfied with their general diet was 3.5% and 4.3% in 2004 and 2014, respectively. For economically insecure participants, the percentage of people not eating balanced meals was 23.7% and 24.8% in 2004 and 2014, respectively. Among the participants who did not have economic insecurity, the percentage of people not eating balanced meals was 17.6% and 16.6% in 2004 and 2014, respectively. For economically insecure participants, the percentage of those poor with self-rated health was 30.1% and 28.4% in 2004 and 2014, respectively. In contrast, for the participants who did not have economic insecurity, the percentage of poor with self-rated health was 17.2% and 14.2% in 2004 and 2014, respectively. These trends were also true when only those aged 75 and older or who had long-term care insurance recipients were included. Essentially, large economic differences were found but few differences by survey year. The only decrease significantly was in self-rated poor health among those without economic insecurity in the aggregate.

**Table 2:** Self-rated health, dietary satisfaction and balanced meals intake for each year by economic status

|  | survey year |  |  |  | p |
| --- | --- | --- | --- | --- | --- |
|  | 2004 |  | 2014 |  |  |
|  | n | % | n | % |  |
| *overall* |  |  |  |  |  |
| dietary satisfaction (n and % of "not satisfied") |  |  |  |  |  |
| have economic Insecurity | 106 | 13.9 | 245 | 16.8 | 0.073 |
| no economic Insecurity | 72 | 3.5 | 95 | 4.3 | 0.149 |
| eat balanced meals (n and % of "no") |  |  |  |  |  |
| have economic Insecurity | 181 | 23.7 | 376 | 24.8 | 0.571 |
| no economic Insecurity | 364 | 17.6 | 372 | 16.6 | 0.397 |
| self-rated health (n and % of "poor") |  |  |  |  |  |
| have economic Insecurity | 230 | 30.1 | 431 | 28.4 | 0.395 |
| no economic Insecurity | 355 | 17.2 | 317 | 14.2 | 0.007 |
| *aged 75 or more* |  |  |  |  |  |
| dietary satisfaction (n and % of "not satisfied") |  |  |  |  |  |
| have economic Insecurity | 18 | 14.3 | 88 | 19.8 | 0.159 |
| no economic Insecurity | 22 | 4.1 | 46 | 5.9 | 0.149 |
| eat balanced meals (n and % of "no") |  |  |  |  |  |
| have economic Insecurity | 28 | 22.2 | 104 | 21.8 | 0.928 |
| no economic Insecurity | 80 | 14.9 | 120 | 14.9 | 0.993 |
| self-rated health (n and % of "poor") |  |  |  |  |  |
| have economic Insecurity | 53 | 42.1 | 213 | 44.7 | 0.590 |
| no economic Insecurity | 148 | 27.5 | 189 | 23.4 | 0.088 |
| *long-term care insurance recipient* |  |  |  |  |  |
| dietary satisfaction (n and % of "not satisfied") |  |  |  |  |  |
| have economic Insecurity | 12 | 25.5 | 45 | 28.1 | 0.726 |
| no economic Insecurity | 14 | 11.5 | 18 | 10.5 | 0.797 |
| eat balanced meals (n and % of "no") |  |  |  |  |  |
| have economic Insecurity | 12 | 25.5 | 35 | 20.1 | 0.421 |
| no economic Insecurity | 20 | 16.4 | 22 | 12.1 | 0.286 |
| self-rated health (n and % of "poor") |  |  |  |  |  |
| have economic Insecurity | 36 | 76.6 | 127 | 73.0 | 0.618 |
| no economic Insecurity | 61 | 50.0 | 95 | 52.2 | 0.707 |

p: chi-square test

### 3.3. Logistic regression analyses

**Table 3** shows the results of the logistic regression analyses for all participants. The p-values for interaction in dietary satisfaction were 0.985 and 0.831 for Models 1 and 2, respectively. The p-values for interaction in balanced meal intake were 0.334 and 0.255 for Models 1 and 2, respectively. The p-values for interaction in self-rated health were 0.261 and 0.300 for Models 1 and 2, respectively. Additionally, the main effect of economic status was large, and the main effect of the year was small for both self-rated health, dietary satisfaction, and balanced meal intake.

**Table 3:** Logistic regression analysis of all participants

|  | Model 1 |  |  |  | Model 2 |  |  |  |
| --- | --- | --- | --- | --- | --- | --- | --- | --- |
| | $\beta$ | SE | OR | 95%CI | $\beta$ | SE | OR | 95%CI |
| dietary satisfaction (ref. "satisfied") |  |  |  |  |  |  |  |  |
| economic insecurity (ref. "no") | 1.496 | 0.159 | 4.466 | (3.268, 6.101) | 1.463 | 0.161 | 4.317 | (3.149, 5.916) |
| year (ref. 2004) | 0.229 | 0.159 | 1.258 | (0.920, 1.718) | 0.132 | 0.161 | 1.141 | (0.832, 1.563) |
| interaction (economic insecurity*year) | -0.004 | 0.203 | p for interaction = 0.985 |  | 0.044 | 0.205 | p for interaction = 0.831 |  |
| intercept | -3.322 | 0.120 |  |  | -3.366 | 0.139 |  |  |
| eat balanced meals (ref. "yes") |  |  |  |  |  |  |  |  |
| economic insecurity (ref. "no") | 0.374 | 0.103 | 1.453 | (1.188, 1.777) | 0.302 | 0.104 | 1.352 | (1.102, 1.658) |
| year (ref. 2004) | -0.069 | 0.081 | 0.934 | (0.796, 1.094) | -0.060 | 0.082 | 0.942 | (0.801, 1.106) |
| interaction (economic insecurity*year) | 0.127 | 0.132 | p for interaction = 0.334 |  | 0.151 | 0.133 | p for interaction = 0.255 |  |
| intercept | -1.544 | 0.058 |  |  | -1.617 | 0.075 |  |  |
| self-rated health (ref. "good/fair") |  |  |  |  |  |  |  |  |
| economic insecurity (ref. "no") | 0.732 | 0.098 | 2.078 | (1.714, 2.518) | 0.851 | 0.101 | 2.342 | (1.921, 2.856) |
| year (ref. 2004) | -0.227 | 0.084 | 0.797 | (0.675, 0.939) | -0.384 | 0.087 | 0.681 | (0.574, 0.807) |
| interaction (economic insecurity*year) | 0.145 | 0.129 | p for interaction = 0.261 |  | 0.137 | 0.132 | p for interaction = 0.300 |  |
| intercept | -1.574 | 0.058 |  |  | -2.123 | 0.081 |  |  |

$\beta$ , logistic regression coefficient; OR, odds ratio; CI, confidence interval.
Model 1: economic status, year, and interaction were used as independent variables. Model 2: Model 1 + gender, age, and living arrangements were adjusted.

**Table 4** shows the results of sensitivity analyses or the logistic regression analyses for the participants “aged 75 or more” and “long-term care insurance recipient.” Both were similar to those analyzed in all participants. All p-values for the interaction were ≥ 0.05.

**Table 4:**
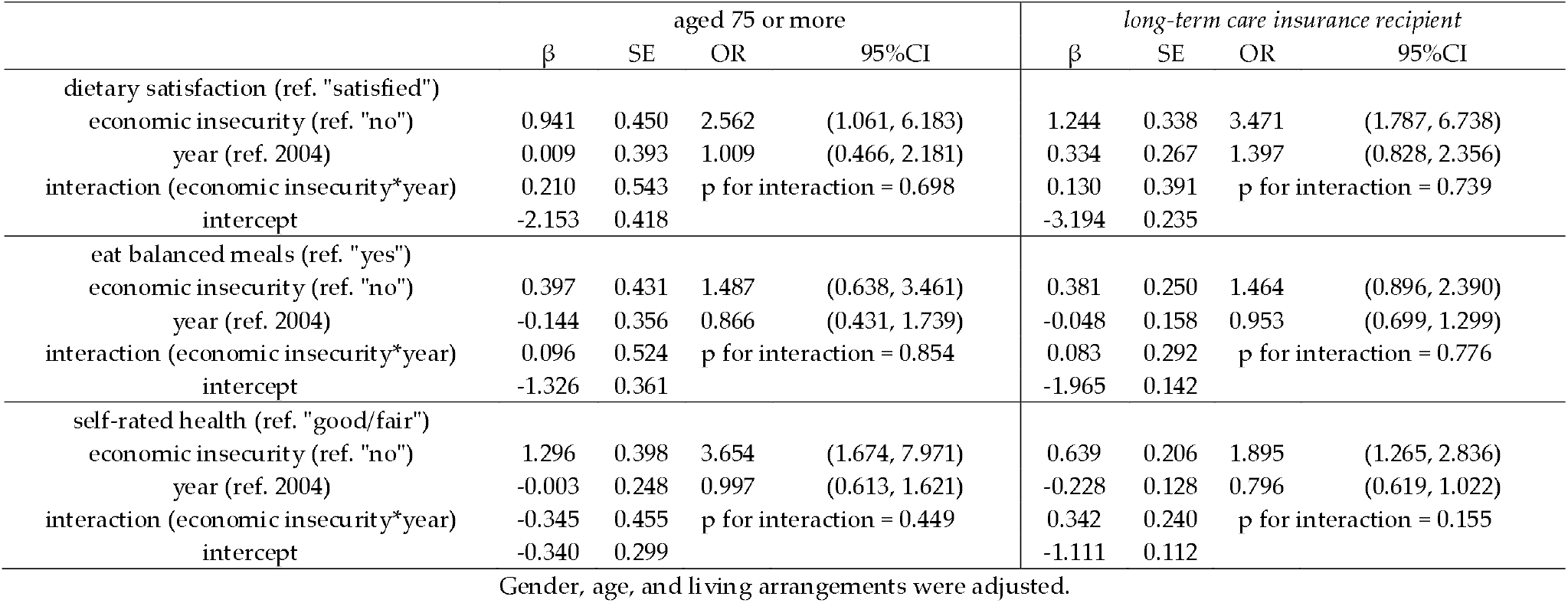
Logistic regression analysis of “aged 75 or more” and “long-term care insurance recipient”

|  | aged 75 or more |  |  |  | long-term care insurance recipient |  |  |  |
| --- | --- | --- | --- | --- | --- | --- | --- | --- |
| | $\beta$ | SE | OR | 95%CI | $\beta$ | SE | OR | 95%CI |
| dietary satisfaction (ref. "satisfied") |  |  |  |  |  |  |  |  |
| economic insecurity (ref. "no") | 0.941 | 0.450 | 2.562 | (1.061, 6.183) | 1.244 | 0.338 | 3.471 | (1.787, 6.738) |
| year (ref. 2004) | 0.009 | 0.393 | 1.009 | (0.466, 2.181) | 0.334 | 0.267 | 1.397 | (0.828, 2.356) |
| interaction (economic insecurity*year) | 0.210 | 0.543 | p for interaction = 0.698 |  | 0.130 | 0.391 | p for interaction = 0.739 |  |
| intercept | -2.153 | 0.418 |  |  | -3.194 | 0.235 |  |  |
| eat balanced meals (ref. "yes") |  |  |  |  |  |  |  |  |
| economic insecurity (ref. "no") | 0.397 | 0.431 | 1.487 | (0.638, 3.461) | 0.381 | 0.250 | 1.464 | (0.896, 2.390) |
| year (ref. 2004) | -0.144 | 0.356 | 0.866 | (0.431, 1.739) | -0.048 | 0.158 | 0.953 | (0.699, 1.299) |
| interaction (economic insecurity*year) | 0.096 | 0.524 | p for interaction = 0.854 |  | 0.083 | 0.292 | p for interaction = 0.776 |  |
| intercept | -1.326 | 0.361 |  |  | -1.965 | 0.142 |  |  |
| self-rated health (ref. "good/fair") |  |  |  |  |  |  |  |  |
| economic insecurity (ref. "no") | 1.296 | 0.398 | 3.654 | (1.674, 7.971) | 0.639 | 0.206 | 1.895 | (1.265, 2.836) |
| year (ref. 2004) | -0.003 | 0.248 | 0.997 | (0.613, 1.621) | -0.228 | 0.128 | 0.796 | (0.619, 1.022) |
| interaction (economic insecurity*year) | -0.345 | 0.455 | p for interaction = 0.449 |  | 0.342 | 0.240 | p for interaction = 0.155 |  |
| intercept | -0.340 | 0.299 |  |  | -1.111 | 0.112 |  |  |

Gender, age, and living arrangements were adjusted.

## 4. Discussion

This study described trends in health and dietary disparities by economic status among the elderly in Japan from 2004 to 2014. As a result, I identified health and dietary disparities by economic status in both 2004 and 2014, but the trends were largely parallel. Comparing 2004 and 2014, the health dietary disparities had not widened, but neither had they narrowed. I believe the results will contribute to future research on health disparities in Japan.

The results of this study show that dietary disparities by economic status remained unchanged from 2004 to 2014. A previous study examined trends in health disparities by economic status among Japan’s elderly from 2004 to 2013 [25]. The results of the previous study [25] remained unchanged as in this study. In another previous study of Japanese adults and elderly, health disparities by economic status widened slightly in 2007 and 2010 compared to 2004, but they were at the same level in 2013 as in 2004 [13]. Results of this study are consistent with those of previous studies [13,25]. This study did not have data for the years between 2004 and 2014 and, thus, could not capture trends in those intermediate years. Additionally, in Japan, the Law for Supporting the Independence of the Indigent was enacted in 2013, and the Independence Support System for the Indigent began operating in 2015 [37]. The support provided by this system may contribute to the improvement of disparities in health and diet. Therefore, analyzing data from 2014 onward in the future.

Additionally, unlike previous studies, this study did not observe a crossover of disparity by age [13,25]. Prior studies observed crossover of health disparities by income around age 80 [13,25]. Essentially, the health disparity by income disappears around age 80, after which an inverse association appears: the lower the income, the healthier the individual. This crossover was not observed for dietary inequalities in the present sensitivity analysis of those aged 75 and older. However, as this study has a small sample size, conducting a more detailed examination of the crossover in dietary disparities by economic status using other data in the future will be necessary.

### Limitations

This study has several limitations. First, this study’s outcomes were subjective items for which validity and reliability were not verified. Validating dietary satisfaction with a validated diet-related quality of life scale, for instance, would be a better option. Dietary balance with nutrient intakes could be calculated based on actual dietary surveys. However, when examining past trends, as in this case, using the best method is often impossible. Therefore, verifying the trend using an indicator such as that used in this study is worthwhile. Moreover, data used in this study were obtained using different survey methods in 2004 and 2014, and the existence of bias due to this difference cannot be denied. Finally, as this is a repeated cross-sectional survey, the same individuals were not followed. While I statistically adjusted for gender, age, and family structure, I cannot eliminate the possibility that I am comparing potentially slightly heterogeneous groups.

## 5. Conclusions

This study examined the trends of health and dietary disparities by economic status among elderly individuals in Japan from 2004 to 2014. The results showed disparities in self-rated health, dietary satisfaction, and intake of a balanced meal due to economic status. Moreover, these disparities remained unchanged from 2004 to 2014. However, this study used data from only two period, 2004 and 2014. Longer-term trends need to be identified in the future.

## Data Availability

Data Availability Statement: It is available by applying to the Social Science Japan Data Archive, Center for Social Research and Data Archive, which is affiliated with the Institute of Social Sciences, University of Tokyo.

## Institutional Review Board Statement

Not applicable. This study was exempt from applying the ethical guidelines for life science and medical research involving human subjects in Japan because anonymous information was derived from a survey conducted prior to the study.

## Informed Consent Statement

Not applicable.

## Data Availability Statement

It is available by applying to the Social Science Japan Data Archive, Center for Social Research and Data Archive, which is affiliated with the Institute of Social Sciences, University of Tokyo.

## Acknowledgments

Data for this secondary analysis, “Survey of Attitudes among the Elderly toward Daily Life” 2004 and 2014, was provided by the Social Science Japan Data Archive, Center for Social Research and Data Archives, Institute of Social Science, University of Tokyo.

